# Inflection in prevalence of SARS-CoV-2 infections missing the N501Y mutation as a marker of rapid Delta (B.1.617.2) lineage expansion in Ontario, Canada

**DOI:** 10.1101/2021.06.22.21259349

**Authors:** Kevin A. Brown, Eugene Joh, Sarah A. Buchan, Nick Daneman, Sharmistha Mishra, Samir Patel, Troy Day

## Abstract

**Background:** The severe acute respiratory syndrome coronavirus 2 (SARS-CoV-2) Delta lineage (B.1.617.2) was implicated in the SARS-CoV-2 surge in India. We sought to describe the rapid expansion of the Delta lineage in Ontario, Canada (population 15 million) using mutation profile information and confirmatory whole genome sequencing.

**Methods:** All laboratory-confirmed SARS-CoV-2 cases reported to Public Health Ontario between April 1^st^ and June 12^th^ 2021, with cycle threshold values ≤35, were eligible for screening for the N501Y and the E484K mutations. We classified cases via mutation screening as: (1) N501Y-/E484K- (wild-type/Delta), (2) Alpha (N501Y+/E484K-), (3) Beta/Gamma (N501Y+/E484K+), or (4) N501Y-/E484K+ (predominantly B.1.525, and B.1.1.318).

**Results:** The N501Y-/E484K- mutation profile went from having a 29% transmission deficit relative to Alpha (relative R_e_ = 0.71, 95%CI: 0.64, 0.77) on April 1^st^ to having a 50% transmission advantage on June 12^th^ (relative R_e_ = 1.50, 95%CI: 1.31, 1.71). Whole genome sequencing of N501Y-/E484K-cases (N=583) confirmed that the pattern of increasing relative reproduction number coincided with the replacement of wild-type with Delta variant (from 2.2% in early April, to 83% in late May).

**Discussion:** Delta is rapidly overtaking other SARS-CoV-2 variants in Ontario, and has a substantial transmission advantage. An inflection in the proportion of cases missing the N501Y mutation from rapidly decreasing to rapidly increasing,^3^ may be an early warning signal for Delta lineage expansion.

## Background

The severe acute respiratory syndrome coronavirus 2 (SARS-CoV-2) Delta lineage (B.1.617.2) was implicated in a devastating SARS-CoV-2 surge in India, and subsequently with increasing rates in the United Kingdom.^1^ The Delta variant lacks the N501Y mutation (possessed by Alpha, Beta and Gamma) and also lacks the vaccine escape mutation E484K (possessed by Beta and Gamma). We use mutation profile information and confirmatory whole genome sequencing to describe the rapid expansion of the Delta lineage in Ontario, Canada (population 15 million) during a period of increasing vaccination (first dose coverage: 13.9% on April 1^st^, 63.4% on June 12^th^),^2^ and estimate the relative transmissibility of the Delta variant.^3^

## Methods

All laboratory-confirmed SARS-CoV-2 cases reported to Public Health Ontario between April 1^st^ and June 12^th^ 2021, with cycle threshold values ≤35, were eligible for N501Y and the E484K mutation screening. We classified cases as: (1) N501Y-/E484K- (wild-type/Delta), (2) Alpha (N501Y+/E484K-), (3) Beta/Gamma (N501Y+/E484K+), or (4) N501Y-/E484K+ (predominantly B.1.525, and B.1.1.318).^4^ To adjust for daily differences in the proportion that received mutation screening, we multiplied the proportion of each profile among those screened, by the total number of cases.

We modeled the daily count of each mutation profile using a count quasi-Poisson regression with the date modeled as a flexible spline (weekly knots) and an interaction term between date and each mutation profile. Daily growth, and the daily proportion of N501Y-/E484K-isolates were derived from this model. Daily growth was translated to a reproduction number (R_e_), assuming a generation interval of 5.2 days.^5^

Because initial confirmed cases of Delta in Ontario were concentrated in the municipality of Brampton, we also stratified analyses across the Greater Toronto Area’s 3 largest municipalities (Toronto [pop. 3,100,000], Mississauga [pop. 900,000], Brampton [pop. 600,000]). Confirmatory whole genome sequencing on a weekly provincial sample of N501Y-/E484K-isolates was used to estimate the proportion that were Delta.

## Results

In total, 185,085 SARS-CoV-2 cases were identified and 153,998 (83.2%) received mutation screening, of which 81.1% were Alpha, 9.6% were N501Y-/E484K-, 5.7% were Beta/Gamma, and 3.6% were N501Y-/E484K+. During the period of rapidly increasing vaccination, the estimated reproduction number of Alpha declined from 1.20 to 0.62 (Figure 1), while that of N501Y-/E484K-increased slightly from 0.85 to 0.93. As such N501Y-/E484K-went from having a 29% transmission deficit relative to Alpha (relative R_e_ = 0.71, 95%CI: 0.64, 0.77) on April 1^st^ to having a 50% transmission advantage on June 12^th^ (relative R_e_ = 1.50, 95%CI: 1.31, 1.71).

**Figure 1.**
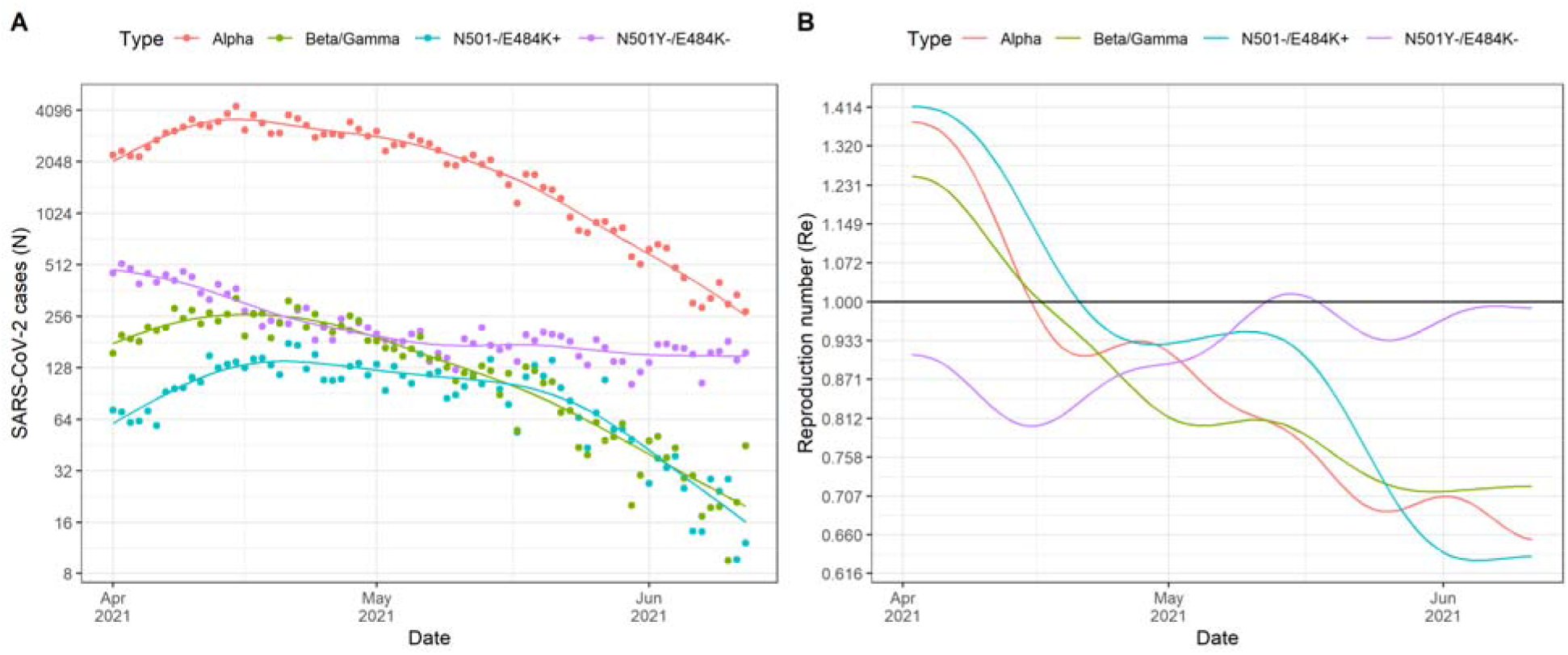
Estimation of the incidence (left panel) and reproduction number (right panel) of mutation profiles in Ontario. Relative to Alpha (red), the reproduction number of the N501Y-/E484K- mutation profile (purple) went from 29% lower than Alpha lineage to 50% higher at the end of the period.

We observed an inflection from a decreasing trend in the proportion of N501Y-/E484K-before April 28, to an increasing trend afterwards (Figure 2A). SARS-CoV-2 cases in the municipality of Brampton (Figure 2B) experienced this inflection first (April 17). Ontario-wide whole genome sequencing on N501Y-/E484K-samples (N=538) revealed an increase in the proportion that were Delta (from 2/87 [2.2%] preceding April 4th, to 10/12 [83%] in the last week of May); in the week of the inflection approximately half of N501Y-/E484K-samples were Delta (53.9%; 41/76).

**Figure 2.**
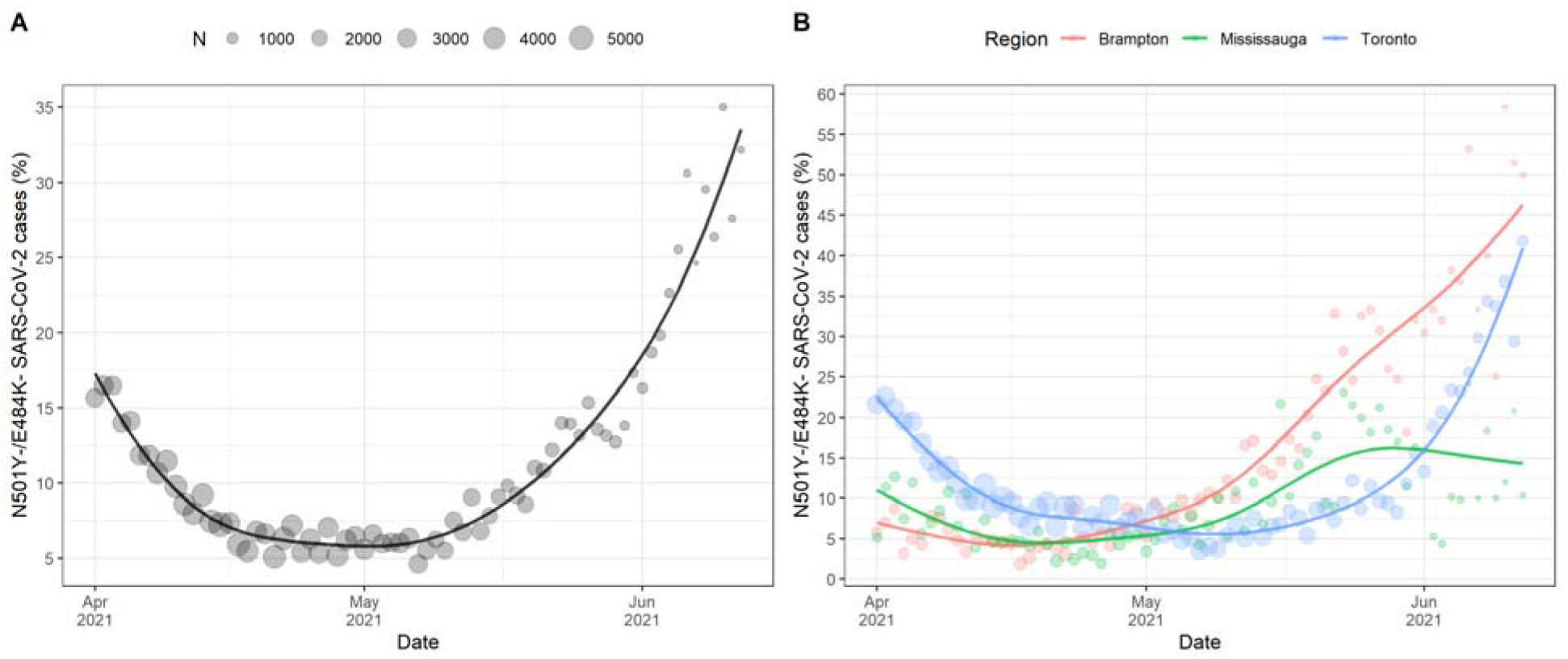
Proportion of SARS-CoV-2 cases with the N501Y-/E484K- mutation profile in Ontario (left panel). We observed an inflection from a rapid decrease in the proportion of N501Y-/E484K- lineages to a rapid increase on April 28. At the end of the study period on June 12^th^, N501Y-/E484K- profile cases represented 34% of SARS-CoV-2 cases in Ontario. SARS-CoV-2 cases in the municipality of Brampton (Figure 2B) experienced this change first (April 17) followed by Mississauga (April 24) and then Toronto (May 7).

## Discussion

This study identified a rapid increase in the proportion of SARS-CoV-2 cases with the N501Y-/E484K- mutation profile in Ontario, demonstrating that Delta was rapidly overtaking the Alpha variant and is 50% more transmissible. Our analyses are consistent with estimates of a 64% secondary attack rate advantage over the Alpha lineage from the United Kingdom.^6^ Limitations include potentially non-representative sampling for whole genome sequencing. For countries once dominated by Alpha, Beta, or Gamma lineages, an inflection in the proportion of cases missing the N501Y mutation from rapidly decreasing to rapidly increasing,^3^ may be an early warning signal for Delta lineage expansion.

## Data Availability

Primary analyses in the manuscript are based on the case counts presented in Figure 1A, and as such analyses can be replicated from this figure.

## Acknowledgements

The authors wish to thank Kirby Cronin for assistance with retrieving and interpreting confirmatory whole genome sequencing data.

